# Clinical assessment and voxel-based morphometry study of untreated Adult Attention deficit hyperkinetic disorders patients

**DOI:** 10.1101/2022.05.28.22271305

**Authors:** Sara Morsy, Sherief Ghozy, Ahmed Morsy, Adam A. Dmytriw, Kevin Kallmas, Sadiq Naveed

## Abstract

**Purpose:** Adult ADHD is one of the most undiagnosed diseases mainly because of the misperception that ADHD is a childhood disease. In this study, we assess the characteristic features of adult ADHD using clinical assessment and structural Magnetic resonance imaging (sMRI)

**Methods:** We obtained structural MRI data from the UCLA Consortium for Neuropsychiatric Phenomics for 21 untreated adult ADHD patients and 21 age and gender propensity-matched control patients. For clinical assessment, we compared the scores of Barrat impulsivity score, Dickman impulsivity inventory II, and Eysenck’s Impulsivity Inventory. We then compared grey matter volume (GMV) between ADHD and control patients using a two-sample t-test. We also assessed the correlation between different clinical assessments and GMV.

**Results:** Based on our results, adult ADHD showed significantly higher impulsivity scores, however, no significant difference in functional impulsivity scores or empathy summary scores. For sMRI, there was a significant decrease of GMV of the left cuneus in female ADHD patients. For clinical assessment scales, only the motor impulsiveness subdomain showed a significant positive correlation with the GMV of the left precuneus.

**Conclusions:** In this study, we assessed the characteristic sMRI features and clinical assessment scores for untreated adult ADHD. Our results show that a study with a bigger sample size can identify diagnostic features for adult ADHD.

## Introduction

Attention deficit hyperkinetic disorder (ADHD) is most commonly thought of as a childhood disorder. It is usually therefore overlooked in adults, and most physicians do not receive adequate training for the diagnosis of ADHD in adults (1). A recent study found that primary care physicians could effectively screen or diagnose adult ADHD, largely based on the presence of two or more symptoms during childhood associated with academic and life difficulties. The adult ADHD prevalence ranges from 1% to 6%, which is considered an underestimation of the disease (1, 2). Most newly diagnosed cases in adults are females and the most predominant symptoms are inattention. The diagnosis in adults is usually overlooked since the absence of a childhood diagnosis often affects the clinician’s decision-making (2). Untreated adult ADHD leads to a higher risk for anxiety, depression, and poor quality of life, and adult ADHD patients have higher inattention and impulsivity, which can cause life difficulty and increased criminal acts (3). Adult ADHD has an inevitable outcome on the quality of life, studies found that diagnosis and treatment of ADHD at a young age has a positive outcome later during adulthood (3). However, most cases with less dysfunctional symptoms and less severe forms of the disease were usually undiagnosed until late. Adding to the difficulty of diagnosis in adults, the high comorbid conditions associated with ADHD (e.g., anxiety, bipolar disorder, and depression) lead to misdiagnosis of the disease.

The difficulty to diagnose ADHD in adults was mainly due to atypical presentation and coping mechanisms. Most scales focus on inattention, impulsivity, and hyperactivity, whereas, in adults, executive dysfunction and higher comorbidity are prominent. A study reported that Adult ADHD patients have higher scores on anxiety and depression scales masking their ADHD symptoms (3). A study based on the World Health Organization (WHO) mental health survey initiative found that most undiagnosed adult ADHD received treatment for other comorbid conditions (4). A review of the literature revealed that even successful treatment of comorbidities did not improve the patient quality of life. On the contrary, adequate treatment of ADHD improved significantly the social and academic performance (5). In addition, adult ADHD patients have developed compensatory mechanisms, e.g., social, organizational, attentional, and motor mechanisms, which make the diagnosis even harder (6).

That’s why in this study, we are investigating the characteristic features of adult ADHD using clinical assessment and MRI studies. Structural MRI (sMRI) analysis has been posited as a method to diagnose mental illnesses. Surprisingly, studies of adult ADHD are rare, as most studies investigate children only or a mixed sample of children and adult ADHD.

sMRI studies of adult ADHD showed characteristics features such as a decrease of GMV in visual areas and left orbitofrontal brain (7, 8). Another study did not find any significant volumetric changes in adult ADHD but only found gender-specific changes of caudate in male patients (9). More studies are needed to characterize untreated and undiagnosed adult ADHD.

Therefore, in this study, we examine the use of sMRI to determine if grey matter voxel size has consistent differences between adult ADHD and control patients.

## Methods

### Participants

The data were obtained from UCLA Consortium for Neuropsychiatric Phenomics dataset. The dataset that was registered in openneuro.org (Accession number: ds000030) provided sMRI with patient characteristics, including diagnosis of ADHD (10, 11). The dataset has structural MRI data of ADHD patients with their corresponding ADHD scales’ results that were used for the analysis. For the control group, we selected age and sex-matched control groups using propensity score matching using R package Matchit (12). The consent for these data is described in the original study (10, 11).

### Structural MRI acquisition and preprocessing

First, we checked the sMRI images for any artifacts, and images were excluded if artifacts were present. The preprocessing steps were undertaken using CAT12: in brief, the images were processed using a denoising filter using spatial adaptive non-local means (SANLM), followed by internal resampling. Then, bias correction and affine registration were completed before the segmentation to enhance the outcome of the standard SPM segmentation.

We performed refined voxel-based processing which includes skull stripping, parcellation of the brain, and detection of white matter hyperintensities that are used during final adaptive maximum a posteriori (AMAP) segmentation and normalization.

The final step in segmentation is AMAP and partial volume estimation, which are considered a precise estimate of the fractional content of each type of the brain per voxel as it does not depend on the prior tissue probability. Next, Diffeomorphic Anatomical Registration Through Exponentiated Lie Algebra (DARTEL) was applied for accurate spatial registration.

After segmentation is done, the segmented images were checked for homogeneity and no images were discarded. Smoothing Gaussian kernel with a full width at half maximum (FWHM) of 8 mm was applied to the segmented images.

### Statistical analysis

Propensity score matching based on age and gender was used to match patients in the control group to those in the ADHD group. Patients’ characteristics were compared between the groups using t-tests if normally distributed or Wilcoxon rank test if otherwise. The analysis was conducted using R statistical language (13).

A comparison of grey matter between ADHD and control groups was performed using a two-sample t-test. Total intracranial volume (TIV) was used as a nuisance factor.

General linear regression was used for the correlation analysis of brain volume of the ADHD group versus scores of the Adult ADHD Clinical Diagnostic Scale, Barratt impulsivity scale, Dickman impulsivity inventory II, and Eysenck’s Impulsivity Inventory with TIV as a nuisance variable. Uncorrected p-value and family-wise correction (FWE) were reported if less than 0.001 and 0.05, respectively.

### Clinical assessment

#### Adult ADHD Clinical Diagnostic Scale (ACDS)

It is a semi-structured scale combining the criteria of ADHD diagnosis in DSM-IV ADHD with criteria observed in adult ADHD (14). The patient has an interview with the clinician where the interview starts with a retrospective assessment of the childhood symptoms of ADHD and then extends the assessment to investigate the symptoms of adult ADHD. In addition to DSM-IV criteria, the clinician also assesses other criteria observed in adult ADHD e.g. difficulty in organization, inattention, … etc (14, 15).

### Barratt impulsivity scale (BIS)

Barratt impulsivity scale (BIS) is a psychometric impulsivity questionnaire used for impulsivity assessment in different diseases. It is considered as one of the gold standard questionnaires for the assessment of the three major impulsivity traits: cognitive, motor, and non-planning; the Likert scale is used to respond to each question (16, 17).

The 30-item BIS-11 is the latest version released in 1995 and it was based on the analysis of the 34-item-BIS-10 responses of 412 undergraduate students, 84 different psychiatric patients, 164 substance abuse, and 73 male prisoners. The scale comprises six subdomains: attention, cognitive instability, motor impulsiveness, perseverance, cognitive complexity, and self-control. However, three main subdomains are usually reported in the cognitive literature, motor, and non-planning. The UCLA study has provided the six scales which allowed for more extensive assessment (16, 17).

### Dickman impulsivity inventory-II

Dickman et al. found impulsivity can be classified into two major classes: dysfunctional and functional impulsivity. This classification separates impulsivity into one with positive outcomes (functional impulsivity) and one with negative outcomes (dysfunctional impulsivity) (18). However, both traits are not correlated with each other and usually, individuals show different scores depending on their personality traits, cognitive functioning, and behavioral outcomes (18, 19). Moreover, sex differences also were reported in many studies (18, 19). Colledani et al. reported that only functional impulsivity scores were higher in men (19), whereas, Claes et al. reported that men had significantly higher functional and dysfunctional impulsivity scores than women (20).

### Eysenck’s Impulsivity Inventory

This scale was designed to assess personality traits of impulsiveness, venturesomeness, and empathy. It is a self-administered 54 items questionnaire that is answered by yes/no. Eysenck et al. assessed the narrow impulsivity which they considered as a sign of psychoticism and venturesomeness which is considered a sign of Extraversion (21).

Comparison between Eysenck and the Dickman impulsivity scale revealed that the dysfunctional impulsivity score is highly correlated with the impulsivity scale of Eysenck, meanwhile, the functional impulsivity showed the strongest correlation with the venturesomeness scale (22).

## Results

### Patient Characteristics

The study included 42 age and gender-matched participants diagnosed with ADHD based on the Diagnostic and Statistical Manual, 5^th^ Edition (DSM).

The race was the only variable that was significantly different between the control and ADHD groups; 19/21 (90.5%) of ADHD patients were white, compared with 15/21 (71.4%) of the control group. There was no significant difference between the participants regarding smoking, religion, school degree, or whether the participants speak one or more languages, Table 1. Comparison between inattentive and impulsive types of ADHD in adults did not reveal any significant difference between the two groups, Table 2.

**Table 2.** Comparison between the predominantly hyperactive/impulsive type to the predominantly inattentive type of ADHD.

|  | level | Predominantly<br>Hyperactive/Impulsive<br>Type | Predominantly<br>Inattentive Type | p |
| --- | --- | --- | --- | --- |
| **n** |  | 5 | 8 |  |
| **Gender (%)** | Female | 1 (20.0) | 3 (37.5) | 0.962 |
|  | Male | 4 (80.0) | 5 (62.5) |  |
| **Age (median [IQR])** |  | 27.00 [26.00, 32.00] | 23.50 [21.75, 26.00] | 0.09 |
| **Cigarette (%)** | current smoker | 1 (20.0) | 1 (12.5) | 0.731 |
|  | No | 2 (40.0) | 5 (62.5) |  |
|  | Past smoker | 2 (40.0) | 2 (25.0) |  |
| **residence (%)** | Alone | 1 (20.0) | 0 ( 0.0) | 0.337 |
|  | In home of parents or children | 1 (20.0) | 0 ( 0.0) |  |
|  | In home of siblings/non-lineal<br>relatives | 0 ( 0.0) | 1 (12.5) |  |
|  | In shared home w/ other<br>relatives/friends | 2 (40.0) | 6 (75.0) |  |
|  | With Partner not married | 1 (20.0) | 1 (12.5) |  |
| **school_fail (%)** | No | 5 (100.0) | 8 (100.0) |  |
| **Marital status (%)** | Separated | 2 (40.0) | 0 ( 0.0) | 0.248 |
|  | Single | 3 (60.0) | 8 (100.0) |  |
| **adopted (%)** | No | 5 (100.0) | 7 (87.5) | 1 |
|  | Yes | 0 ( 0.0) | 1 (12.5) |  |
| **sexuality (%)** | Bisexual | 1 (20.0) | 1 (12.5) | 0.928 |
|  | Heterosexual | 3 (60.0) | 5 (62.5) |  |
|  | Homosexual | 1 (20.0) | 2 (25.0) |  |
| **religion (%)** | Catholic | 2 (40.0) | 3 (37.5) | 0.661 |
|  | Jewish | 0 (0.0) | 1 (12.5) |  |
|  | Not Affiliated | 0 (0.0) | 1 (12.5) |  |
|  | Protestant | 3 (60.0) | 3 (37.5) |  |
| **School years (median [IQR])** |  | 16.00 [13.00, 16.00] | 14.50 [14.00, 15.00] | 0.652 |
| **School degree (%)** | Associate's degree | 0 (0.0) | 1 (12.5) | 0.362 |
|  | Bachelor's degree | 3 (60.0) | 1 (12.5) |  |
|  | High school | 1 (20.0) | 1 (12.5) |  |
|  | Other | 0 (0.0) | 1 (12.5) |  |
|  | Some college | 1 (20.0) | 4 (50.0) |  |
| **Children number (median [IQR])** |  | 0.00 [0.00, 0.00] | 0.00 [0.00, 0.00] | 0.206 |
| **Language (%)** | English | 4 (80.0) | 7 (87.5) | 0.325 |
|  | Other | 0 (0.0) | 1 (12.5) |  |
|  | Spanish | 1 (20.0) | 0 (0.0) |  |

Voxel-based morphometry comparing between ADHD and the control group revealed an increase in the volume of the left fusiform in the control group (MNI coordinates = -60, -41, -26, k = 336 voxels, uncorrected p-value = 0.001), however, it was non-significant at FWE<0.05 Figure 1. Moreover, in female adult ADHD patients, there was decreased GMV of left cuneus (MNI coordinate = -11, -81, 24), FWE = 0.02, Figure 2. Worth mentioning, there was no significant difference in the gray matter in the predominantly hyperactive/impulsive type to predominantly inattentive ADHD.

**Figure 1.**
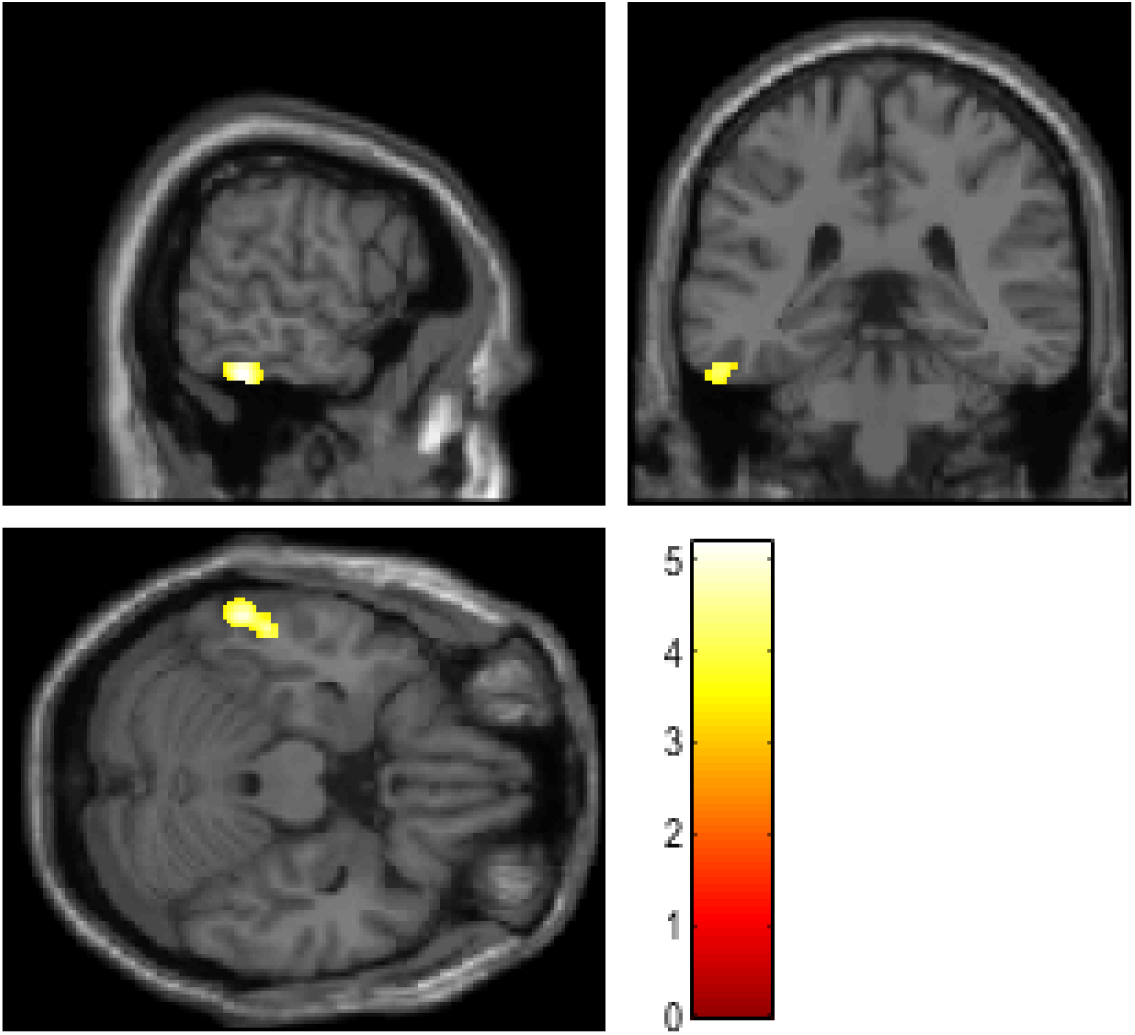
A significant cluster of increased gray matter volume of left fusiform in the control group compared to untreated adult ADHD

**Figure 2.**
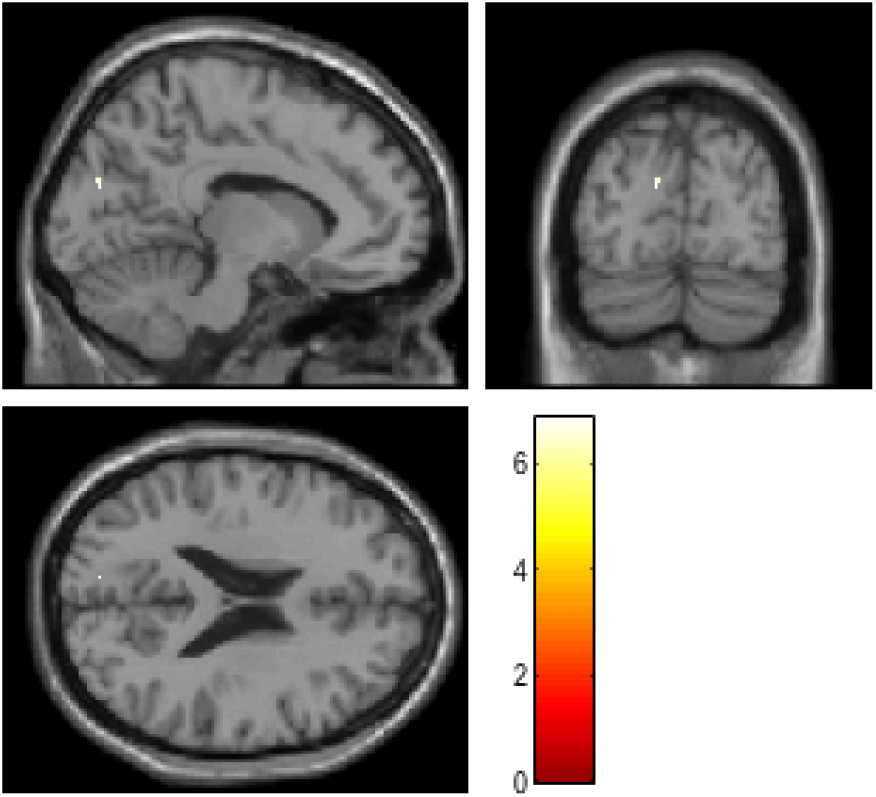
Region of significant decreased GMV of the left cuneus in untreated adult female ADHD compared to the control group.

### Clinical assessment

Four clinical assessment questionnaires were used to evaluate the patients: Adult ADHD Clinical Diagnostic Scale, Barrat impulsivity score, Eysenck’s Impulsivity Inventory, and Dickman Impulsivity Inventory. Statistical analysis revealed that there was a significant difference of all domains of Barrat impulsivity scores between ADHD and control groups, but none of these domains showed a significant correlation with gray matter size, Table 3. For Eysenck’s Impulsivity Inventory, all domains showed significant differences except the empathy summary score. For Dickman impulsivity inventory, only dysfunctional impulsivity scores showed a significant difference between ADHD and control.

**Table 3.**
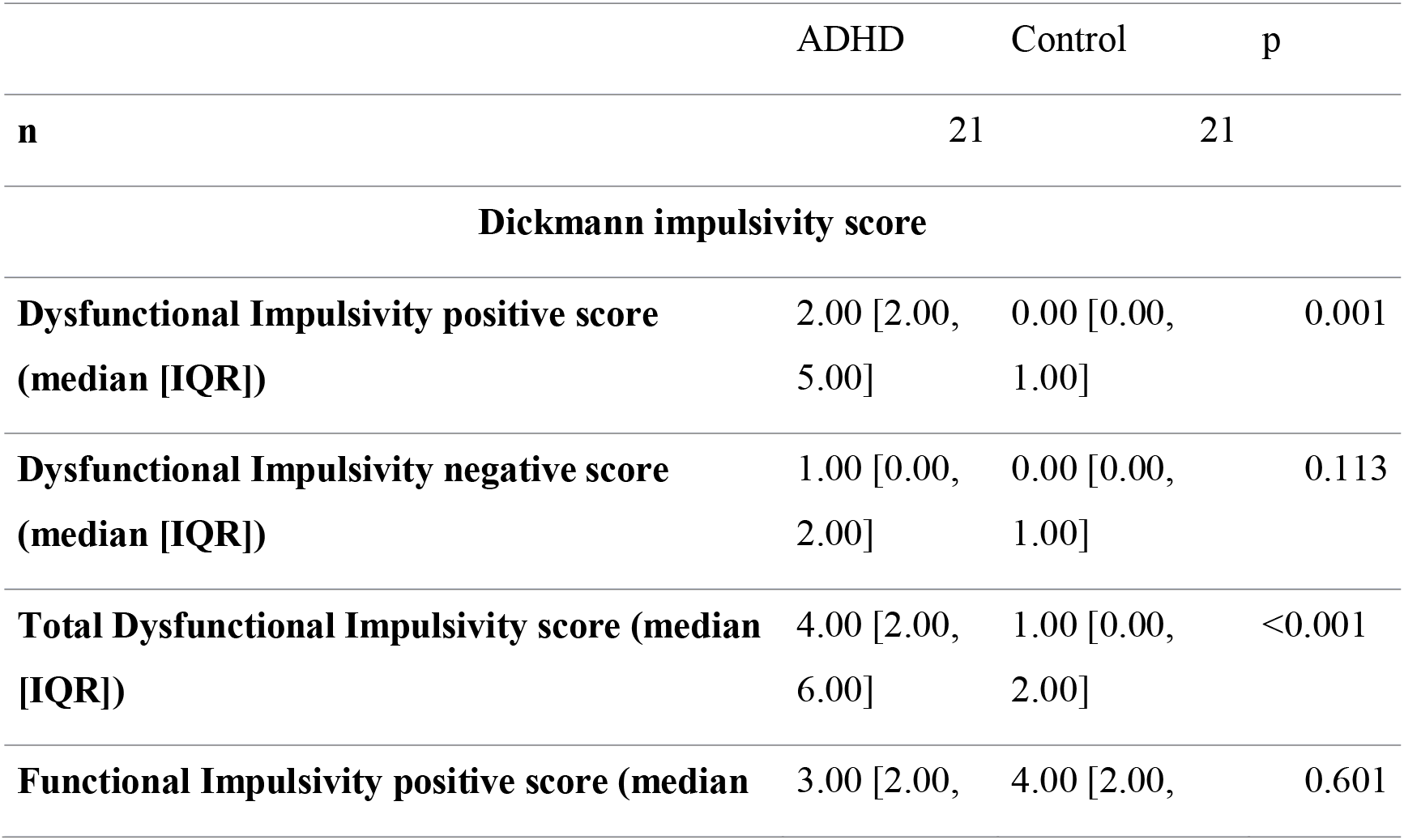
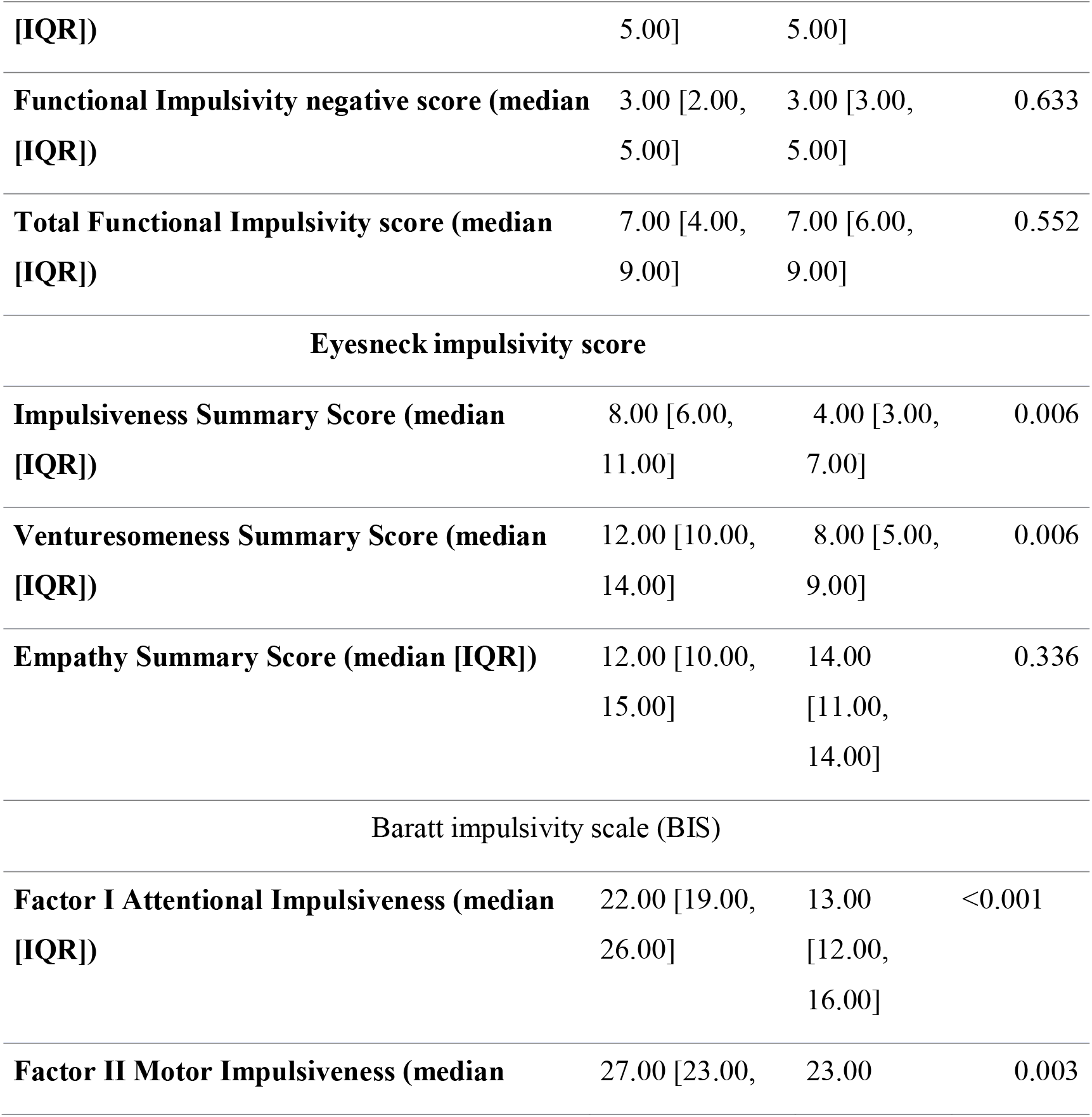
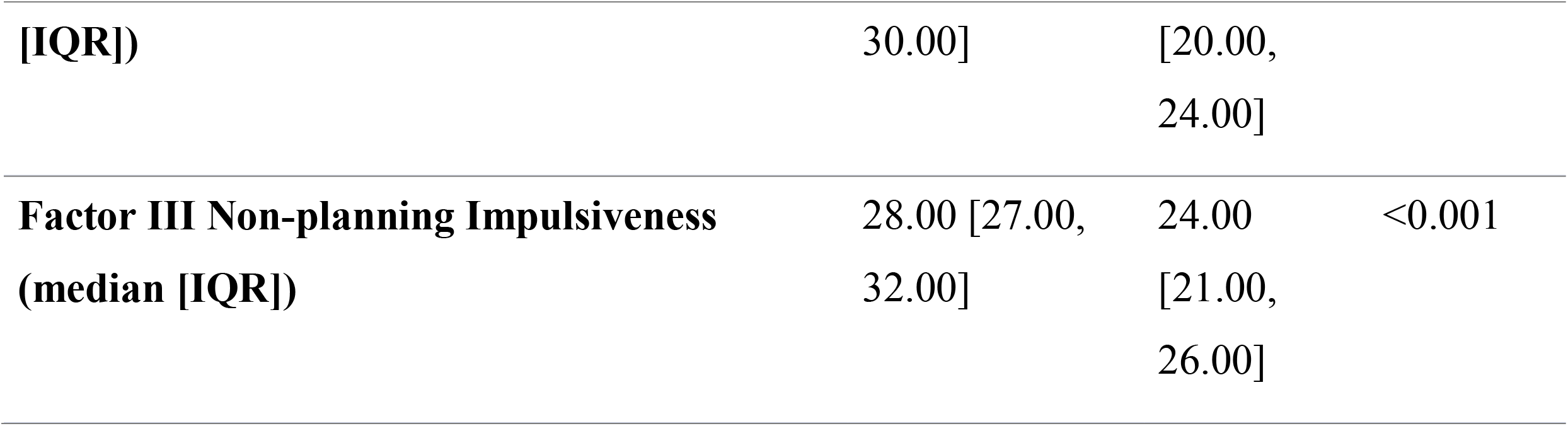
Comparison of clinical assessment between the adult ADHD and control group.

Voxel-based morphometry revealed a positive correlation between the motor impulsiveness subdomain of BIS and GMV of the left precuneus, Figure 3. Other clinical assessment tests did not show any significant correlation with gray matter volume Table 4.

**Figure 3.**
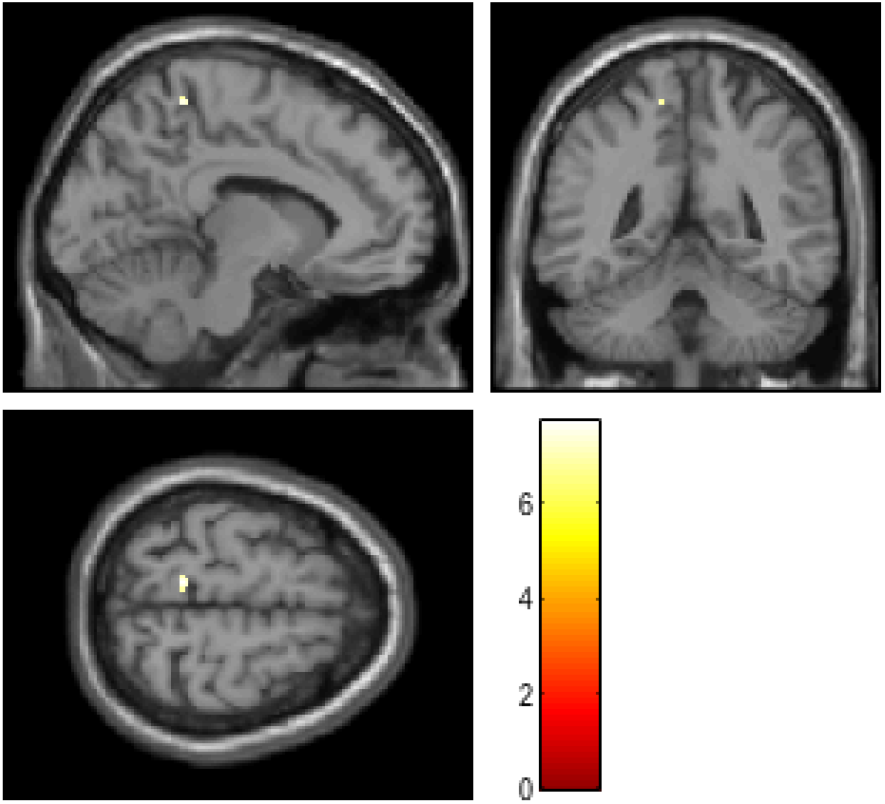
Left precuneus GMV is positively correlated with the motor impulsiveness subdomain of BIS in untreated adult ADHD.

## Discussion

In this study, our voxel-based morphometry (VBM) analysis reveals characteristic sMRI features that can be used for assessing adult ADHD. Gray matter volume (GMV) of the Left Fusiform gyrus was decreased in ADHD adult patients. In addition, our results revealed that adult female ADHD patients had decreased GMV of the cuneus. For clinical assessment scales, only the left precuneus showed a significant positive correlation with the motor impulsiveness scores of BIS. Furthermore, all clinical assessment scores were significantly different between adult ADHD and control group except the functional impulsivity scores of Dickmann impulsivity and empathy summary scores of Eyesneck impulsivity score.

Compared to the control group, we found a significant decrease in the left fusiform gyrus GMV in ADHD patients, nevertheless, this significance did not persist at <0.05. Similar to our findings, under activation and reduction of the GMV was observed among ADHD patients, compared to healthy controls, at the level of the fusiform gyrus in some general population studies (23-25). A previous microstructural study showed an increased radial diffusivity in the right fusiform gyrus of the combined ADHD subtype, which is linked to myelination and GMV (26, 27). Another analysis of electroencephalography, using low-resolution electromagnetic tomography, found increased connectivity between the left middle frontal gyrus and the fusiform gyrus, suggesting the fusiform gyrus’s role in ADHD pathogenesis. Furthermore, studies contradicting our findings have reported the “right” fusiform gyrus, whereas a functional dissociation between both sides was concluded earlier, with a reported possible dominance of the left hemisphere in face perception (identified impairment in ADHD children) (28-30). In a recent VBM meta-analysis, ADHD patients showed a reduced GMV in the ventromedial orbitofrontal cortex (vmOFC) compared to healthy controls. However, no significant differences were reported in comparing the GMV of both groups at the level of the fusiform gyrus (31). The same findings of reduced GMV in vmOFC of ADHD patients were reported in previous meta-analyses as well as multiple large-scale studies, again with no reference to any differences at the fusiform gyrus level (32-36).

Our study showed that there is decreased GMV of the left cuneus in female ADHD patients compared to the female control group (FEW<0.05). There was a GMV difference between male and female ADHD, however, it was not significant at FWE<0.05. Worth mentioning, Cuneus is implicated in the pathogenesis of ADHD which is involved in default mode network and vision as evidenced by the fMRI study which explains its correlation with inattentive scores (37). Another study found increased connectivity in the left cuneus in ADHD patients during working memory tasks (38). Another diffusion tensor imaging study reported increased radial diffusivity of the left cuneus in inattentive ADHD compared with the control group, which indicates myelination deficits in the cuneus (26, 39). Moreover, a study found a significant correlation of cuneus with inattentive scores in ADHD patients (40). In accordance with our results, fMRI studies revealed that there was a significant difference in connectivity between adult female and male ADHD patients in the middle frontal gyrus and cuneus (41, 42). Another study reported thinning of vortex in the cuneus compared to the control group, however, no gender effect was investigated (43). Decreased cuneus GMV is reported in many studies which seemed a characteristic feature of adult ADHD, however, in our study, it was only specific to female ADHD compared to the control group (31).

One of our findings was increased left anterior cingulum and right med frontal orbital gyrus in male ADHD compared to female patients; these regions are responsible for the execution, alerting, and attention (44). These results support the evidence that female ADHD is predominantly inattentive compared to male ADHD which is predominantly impulsive (45). For clinical assessment scores, our results showed higher impulsivity scores for ADHD patients compared to the control group except for the functional impulsivity scores. This explains the reasons for adults with ADHD being unnoticed or considered asymptomatic due to the coping mechanisms that adults develop to compensate for dysfunctional impulsiveness (6).

Furthermore, our study showed an insignificant difference in the Empathy Summary Score between adult ADHD and the control group. This is supported by findings that ADHD shows less emotional empathy but similar cognitive empathy to the control group (46). Cognitive empathy was assessed using the Eyesneck impulsivity score, which explains our results (47). Our results are consistent with another study that found that there was no significant difference in empathy between ADHD and the control group. In addition, they found that empathy was significantly correlated with adulthood symptoms, not childhood symptoms. Moreover, left precuneus GMV significantly correlated with motor impulsiveness of BIS. Meta-analysis of MRI revealed significant involvement of the precuneus in the pathogenesis of ADHD. fMRI showed that there is an overactivation of connectivity networks during cognitive tasks (31). Precuneus is involved in spatial orientation which may explain its relation to motor impulsiveness, in addition, part of the precuneus called “Pep” is involved in visual distraction and attention impairment in ADHD (48, 49).

### Limitations

Our main limitation was the small sample size which affected the power of our study. In addition, we could not do longitudinal MRI analysis to track gray matter changes from childhood to adulthood due to the absence of these data. Future studies should address these changes with a larger sample size.

## Conclusion

In this study, clinical assessment for adult ADHD showed significantly higher dysfunctional impulsivity, inattention, and motor impulsiveness than the control group. On the other hand, sMRI analysis showed consistent results with literature as Fusiform gyrus GMV decreased in ADHD patients. In the female subgroup, cuneus was the predominant brain structure that was significantly associated with the inattentive trait. However, our results should be interpreted carefully due to the small sample size.

## Data Availability

The data that support the findings of this study are openly available in [openneuro.org], reference number [Accession number: ds000030].

https://openneuro.org/datasets/ds000030/versions/00016

## Authors ‘Contributions

SM accounted for the study idea and design, acquisition of data from the openneuro.org database, statistical analysis, and interpretation of data. SM, SG, AM, KK, AD, SN contributed to writing the manuscript. All authors read and approved the final version of the manuscript.

## Acknowledgments

None

## Data availability

This data was obtained from the Openneuro database. Its accession number is ds000030 https://openneuro.org/datasets/ds000030/versions/00016

## Participants consent

These data is acquired from Openneuro database, the patients’ consent for these data is described in the original study (10, 11)

